# Correlation of Body Mass Index (BMI), initial neutralizing antibodies (nAb), ABO group and kinetics of nAb and anti-nucleocapsid (NP) SARS-CoV-2 antibodies in convalescent plasma (CCP) donors – A longitudinal study with proposals for better quality of CCP collections

**DOI:** 10.1101/2020.11.12.20230391

**Authors:** S Wendel, R Fontão-Wendel, R Fachini, G Candelaria, P Scuracchio, R Achkar, MA Brito, LFL Reis, A Camargo, M Amano, RRG Machado, D Araujo, CP Soares, E Durigon

## Abstract

**Introduction:** A cohort of COVID-19 convalescent volunteers allowed the study of neutralizing (nAb) and ligand antibodies kinetics by providing sequential samples during a median of 100 days after onset of disease.

**Material and Methods:** A cohort of previously RT-PCR+ve (detected by nasopharyngeal swab during the acute phase), male convalescent patients, all with mild symptoms, were enrolled on serial blood sample collection for evaluation of longitudinal nAb titers and anti-nucleocapsid (NP) antibodies (IgM, IgG and IgA). Nabs were detected by a cytopathic effect-based virus neutralization test (CPE-based VNT), carried out with SARS-CoV-2 (GenBank: MT350282)

**Results:** A total of 78 male volunteers provided 316 samples, spanning a total of 4820 days of study. Although only 25% of donors kept nAb titers ≥160, after a median of 100 days after the onset of disease, there was a high probability of sustaining nAB titers ≥160 in volunteers whose initial nAb titer was ≥1280, weight ≥ 90kg or BMI classified as overweight or obese, evidenced by Kaplan-Meier estimates and Cox hazard regression. There was no correlation between ABO group, ABO antibody titers and persistent high nAb titers. High IgG anti-NP (S/CO ≥5.0) is a good surrogate for detecting nAB ≥160, defined by ROC curve (sensitivity = 90.5%; CI95% 84.5-94.7%)

**Conclusion:** Selection of CCP donors for multiple collections based on initial high nAb titers (≥1280) or overweight/obese (BMI) provides a simple strategy to achieve higher quality in CCP programs. High IgG anti-NP levels can also be used as surrogate markers for high nAb screening.

## Introduction

COVID-19 convalescent plasma (CCP) has been used for therapy in severe affected COVID-19 patients ^1,2,3,4,5^. The rational relies on the presence of nAb in convalescent’s bloodstream, suppressing patient’s viremia ^6,7^. Several therapeutic approaches have been proposed for COVID-19 treatment, most of them failing under RCTs ^8,9,10^, except for dexamethasone ^11,12^. Despite the controversial results with CCP, the procedure is safe and it still remains one of the few specific therapeutic approaches currently available ^13^. Experience has been gained, but nevertheless the source of CPP is limited and not all suitable donor candidates meet the defined requirements for donation. Thus, until definitive, large scale medical strategies either for prevention (vaccines) or treatment (e.g, cocktail of monoclonal antibodies) ^14^ are effectively available, we consider to be important to define potential additional strategies that would increase the quality of CPP collection.

Little is known about the neutralizing (nAb) and anti-nucleocapsid (NP) antibodies (IgM, IgG and IgA) in CCP donors. From our previous published cohort ^15^, we have detected a positive correlation between donor’s weight and nAb titers. In addition, the ABO group correlates with disease severity (O as protector) ^16^. Since one of the supposed pathogenic cause of severe COVID is the extreme host’s immune response (related to the initial nAb titer produced by the patient), we studied the role of donor’s weight, body mass index (BMI), ABO type and the nAb titer measured at the first sample collection concerning the kinetics of nAb and ligand anti-NP, to define whether there is a special group of CCP donors that could sustain a high and prolonged (&[gt]100 days after onset of disease) level of nAb, resulting in a potential, more efficient CCP program.

### Material and Methods

A cohort of previously RT-PCR+ve (nasopharyngeal swab), male convalescent patients (age 18-60 years, body weight &[gt]55kgs, and full clinical recovery ≥ 14 days, all with mild symptoms), were enrolled on serial blood sample collection for evaluation of longitudinal nAb titers and anti-NP antibodies (IgM, IgG and IgA). This was part of a major cohort for a CCP plasma donation program ^15^. Procedures were carried out according to national legislation ^17, 18^. Individuals were invited for a longitudinal kinetics study, with an intended 100-day period after onset of disease, with an average 7-day collection interval. The initial onset of symptoms was based on description by participants at their first medical screening interview. Approved participants signed an informed consent form, and underwent a series of tests:

Nabs were detected by a cytopathic effect-based virus neutralization test (CPE-based VNT), carried out with SARS-CoV-2 (GenBank: MT350282) in 96-well plates containing 5×10^4^cells/mL of Vero cells (ATCC CCL-81) ^19^, performed in a biosafety level 3 laboratory (BSL-3). NAb titers were transformed in natural logarithm (ln) for normal distribution. Specific IgM, IgG and IgA anti-NP antibodies were detected by ELISA ^20^, using polystyrene plates coated with the nCoV-PS-Ag7 nucleoprotein antigen (Fapon Biotech Inc., Dongguan, China). Results were displayed as the optical density (OD) at 450nm.

All participants were tested for ABO and RhD, irregular antibodies to red blood cell antigens (immunohematological tests); ABO antibody titers (anti-A, anti-B and anti-A,B) were performed at room temperature (RT) and by the human antiglobulin test (AHG) using an automated platform (IH-1000 platform - Biorad, Switzerland). No anti-HLA antibodies were tested (male individuals).

Variable distributions were checked by the Shapiro-Wilk tests. Comparisons were made by t-test, Fisher exact test, ANOVA, two-tailed Mann-Whitney U test, two-tailed Spearman’s correlation, Wilcoxon matched-pairs signed-rank or Kruskall-Wallis test. Bonferroni adjustment was employed whenever possible. The Kaplan-Meier test equality for outcome was evaluated by the two-sided log rank. As outcome, we defined a total of 5 consecutive collection or if a given sample collection had a nAb titer ≤80. Hazard ratio was evaluated by the Cox hazard regression. Values with p &[lt]0.05 were considered significant. Tests and figures were made by STATA v15 (College Station, Tx) and JMP 15.2.1 (Cary, NC) statistical packages.

IRB approval - Study was approved by our IRB and the Brazilian Commission on Ethics and Research (CONEP) under request CAAE: 30259220.4.2001.5461.

## Results

There were initially a total of 149 candidates who participated in the original cohort and had their first sample collected. The initial data displayed a total of 15/149 (10%) cases belonging both to the heavy weight (≥ 91kg) and high nAb titers (≥ 1280) group, with 11/15 (73.3%) belonging to A or AB groups and only 4 from O group. From this initial data, we correlated whether ABO groups, divided either into A-type (A/AB) and non-A (O/B) or O and non-O groups had some role in the maintenance of high nAb. Our initial nAb distribution, according to weight (kg)/BMI (kg/m^2^) and A-type (A x non-A) are shown in Figure 1.

**Figure 1.**
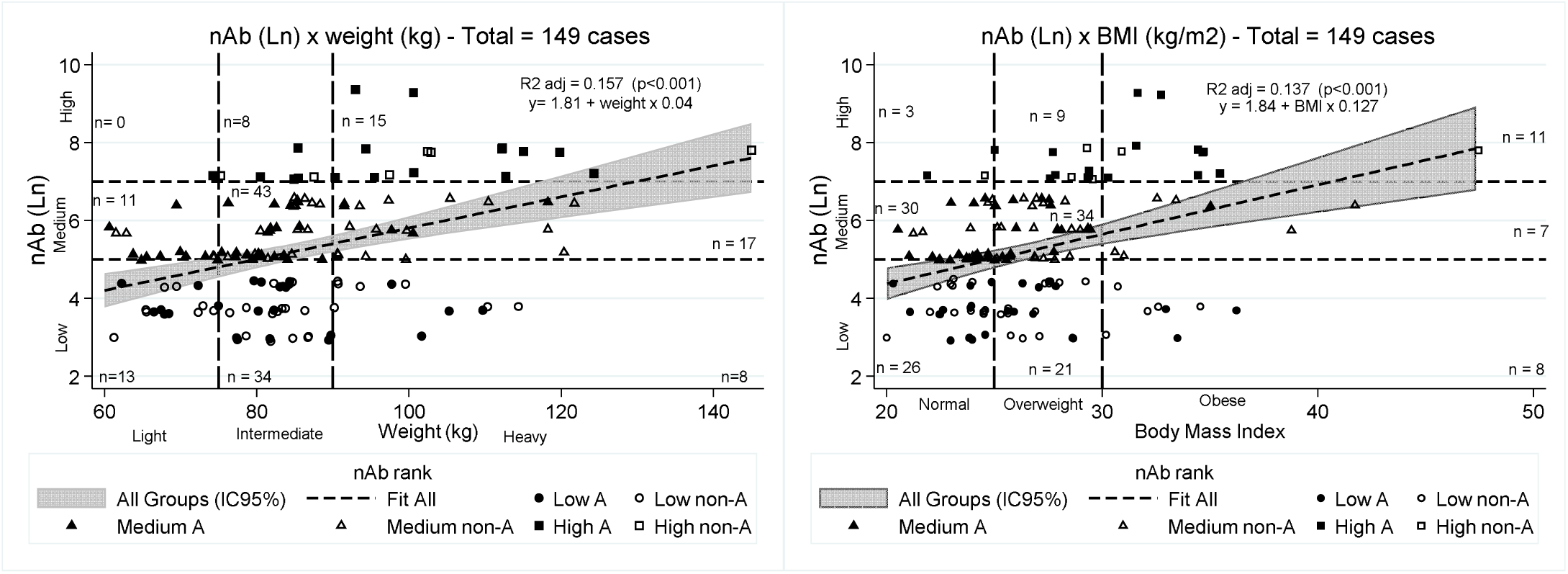
Initial nAb titer distribution based on weight (***left***) and BMI (***right***) from the initial 149 potential candidates in the study. Due to a striking difference in the initial nAb titers, we ranked the cohort into three (3) main titer categories: ***low*** (nAb ≤80); ***medium*** (640≤ nAb≥160); and ***high*** (nAb ≥1280). A major prevalence of A-type individuals (11/15; 73.3%) both from the higher nAb and heavy weight groups was observed, which led us to focus our evaluation on ABO group during the sequential collections.

From the initial 149 individuals, only 78 volunteers provided at least two serial samples, enabling to perform the longitudinal kinetics study. As control group for ABO tests, we evaluated 87 first-time blood donors who donated blood before the arrival of COVID-19 pandemic in SaoPaulo (A=30, O=30 and B=27 cases; no AB was included). The demographics summary is shown in Table 1 and S1 (supplement). This cohort was capable of providing a total of 316 blood samples (up to 5 serial collections) for nAbs titers and Anti-NP, shown in Table 2.

**Table 1.** General main demographics (age, weight, height and body mass index – BMI) of 87 first-time donors for ABO control (right) and 78 male volunteers for the nAb study (left).

|  | Control (n=87) | Cohort (n=78) |  |  |  |  |  |
| --- | --- | --- | --- | --- | --- | --- | --- |
| | Mean ± sd | Range | Median | Mean ± sd | Range | Median | P value |
| Age (years) | 33.3 ± 9.6; | 20.0-55.0 | 33.0 | 36.7 ± 9.1 | 21.0-60.0 | 36.5 | 0.0212 |
| Weight (kg) | 85.2 ± 13.1 | 65.0-127.0 | 83.0 | 86.8 ± 15.1 | 60.0-124.0 | 85.0 | 0.4672 |
| Height (m) | 1.8 ± 0.1 | 1.7-2.0 | 1.8 | 1.8 ± 0.1 | 1.7-2.0 | 1.8 | 1.000 |
| BMI (kg/m<sup>2</sup>) | 26.8 ± 3.6 | 20.0-38.3 | 25.9 | 27.1 ± 4.1 | 20.0-41.9 | 26.8 | 0.6174 |
|  |  |  |  | BMI (kg/m<sup>2</sup>) | N | % |  |
|  |  |  |  | Normal | 31 | 39.7 |  |
|  |  |  |  | Overweight | 33 | 42.3 |  |
|  |  |  |  | Obese | 14 | 18.0 |  |

**Table 2.**
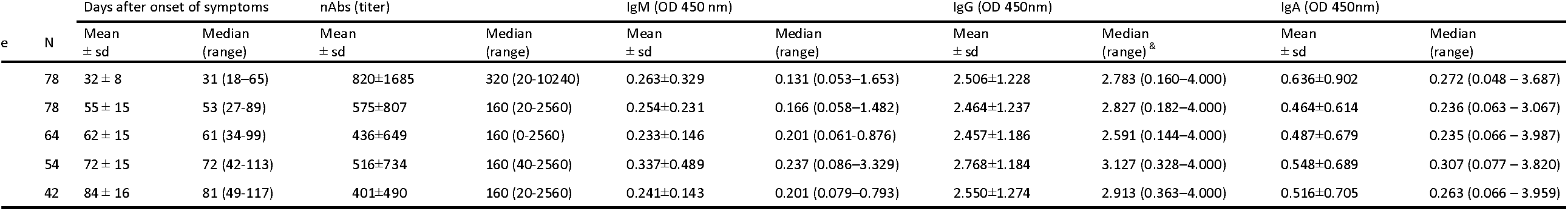
Evaluation of 316 blood sample collections from 78 male volunteers, measured as days after onset of symptoms, nAb titers, IgM, IgG and IgA levels (OD 450nm). There is a trend for nAb titer declining after serial collections (Wilcoxon sign rank test, p&[lt]0.001). Statistical significance among serial samples (Wilcoxon sign rank test) was observed for both the initial nAb (titer) and all subsequent collections, and also between nAb collection #2 x collections #3 and #5. All other results, including for IgG and IgM showed no statistical significance. Legends:^&^ Maximum OD (450 nm)

Figures 2, S1 and S2 (supplement) show that nAb had a different behavior, divided into three ranks: low (nAb ≤80); medium (640≤ nAb≥160); and high (nAb ≥1280). The parallel plot shows that those with initial low titers (Group 1; n=12) kept a sustained low nAb titer, preventing participants from future plasma donation. Group 2 (medium; n=49) had a modest decline in nAb titers, though most participants sustained levels high enough for donation. Group 3 (high, n=17), despite showing the most persistent nAb titer decline, kept their levels above the minimum necessary for plasma donation (≥160), being the best candidates for a CCP program. The heat map gives an overall impression of the nAb titer kinetics.

**Figure 2.**
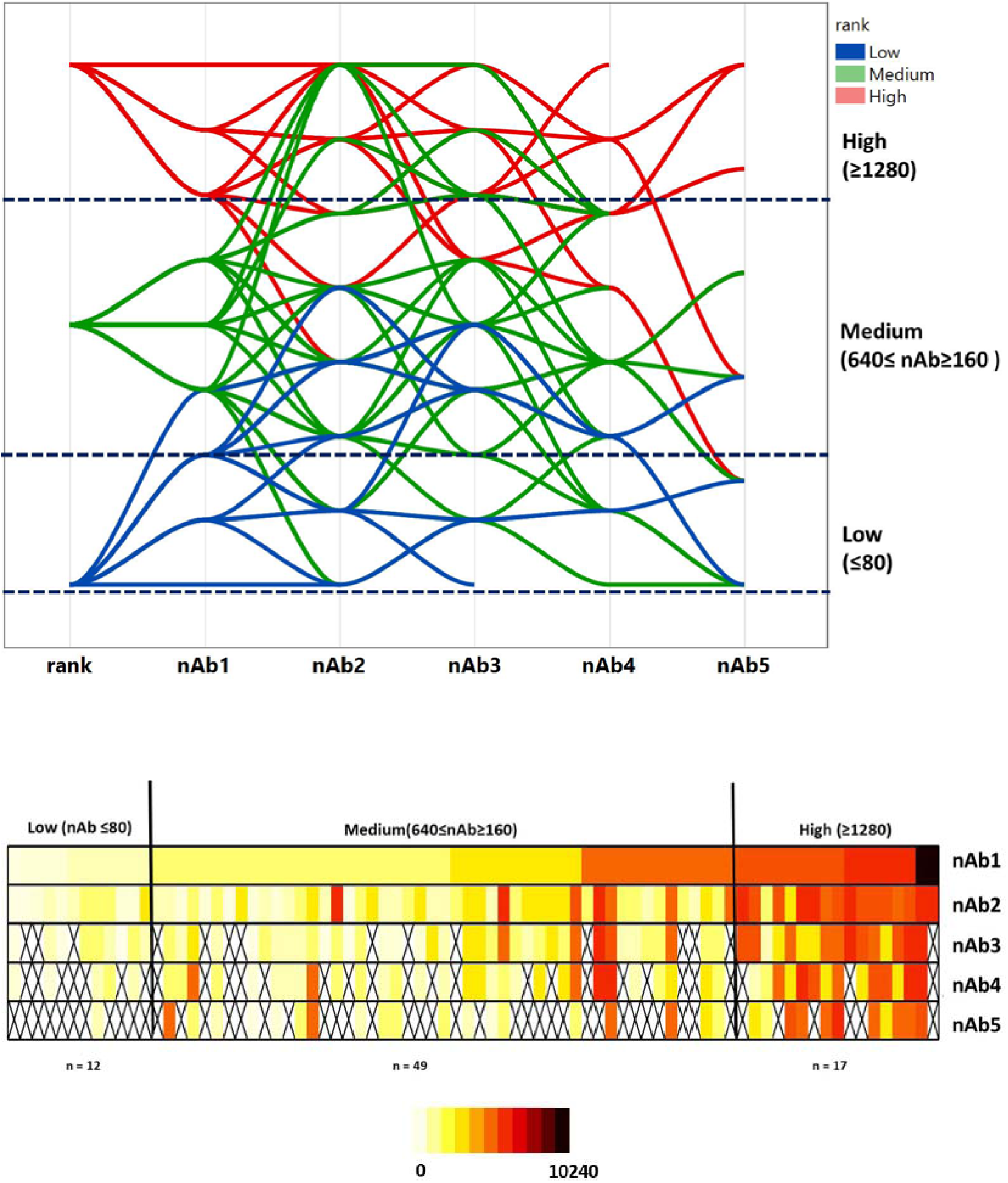
Neutralizing antibodies (nAb) of cohort (n=78 individuals; 316 collections). ***Top:*** Parallel plot of sequential samples, based on the initial nAb titer (nAb 1, 2, 3, 4, 5 rank). Group 1 (low, blue line; n=12) kept a sustained low nAb titer, which prevented the participants from future plasma donation. Group 2 (medium, green line; n=49) had a modest decline in nAb titers, though most of participants sustained levels high enough for donation. Group 3 (high, red line; n=17), despite showing the most persistent nAb titer decline, always kept their levels above the threshold for plasma donation (≥160), being the best candidates for a CCP program. ***Bottom:*** Heat map of all consecutive samples.

For anti-NP (IgM, IgG and IgA), we have transformed the absorption (DO 450nm) by the correspondent cut-off (0.2 - IgM/ IgA; 0.3 - IgG), enabling comparable results for multiple tests. Because the S/CO ratio followed a non-Gaussian distribution, it was transformed into natural logarithm (Ln). Figure 3 shows that except for IgM, there was no statistical significance for the whole group between the initial and all subsequent collections (Wilcoxon sign rank test). In our initial work on CCP ^15^, we have demonstrated that IgG S/CO levels ≥ 5 detected 82.4% of samples with nAb titer ≥ 160. There was again a very good correlation between high IgG levels (S/CO ≥5 and nAb titer ≥ 160), as shown in Figure 4. The adjusted R^2^ for the five sets of collection was very stable (from 0.2199 to 0.3970; all p ≤ 0.001). An ROC curve demonstrated this cut-off had a sensitivity for detecting samples with nAb titers ≥160 of 90.5% (CI95%: 84.5 – 94.7%), specificity of 41.8% (CI95%: 34.8 – 49.17%), and positive and negative likelihood ratio (LR) of 1.556 and 0.2276, respectively. The specificity of the test shouldn’t be considered a major problem, once all individuals were allegedly infected by SARS-CoV-2. The contour plot with the corresponding cluster of 10-density levels, reveals how concentrated are the high IgG S/CO samples, showing a good correlation with high nAb titers.

**Figure 3.**
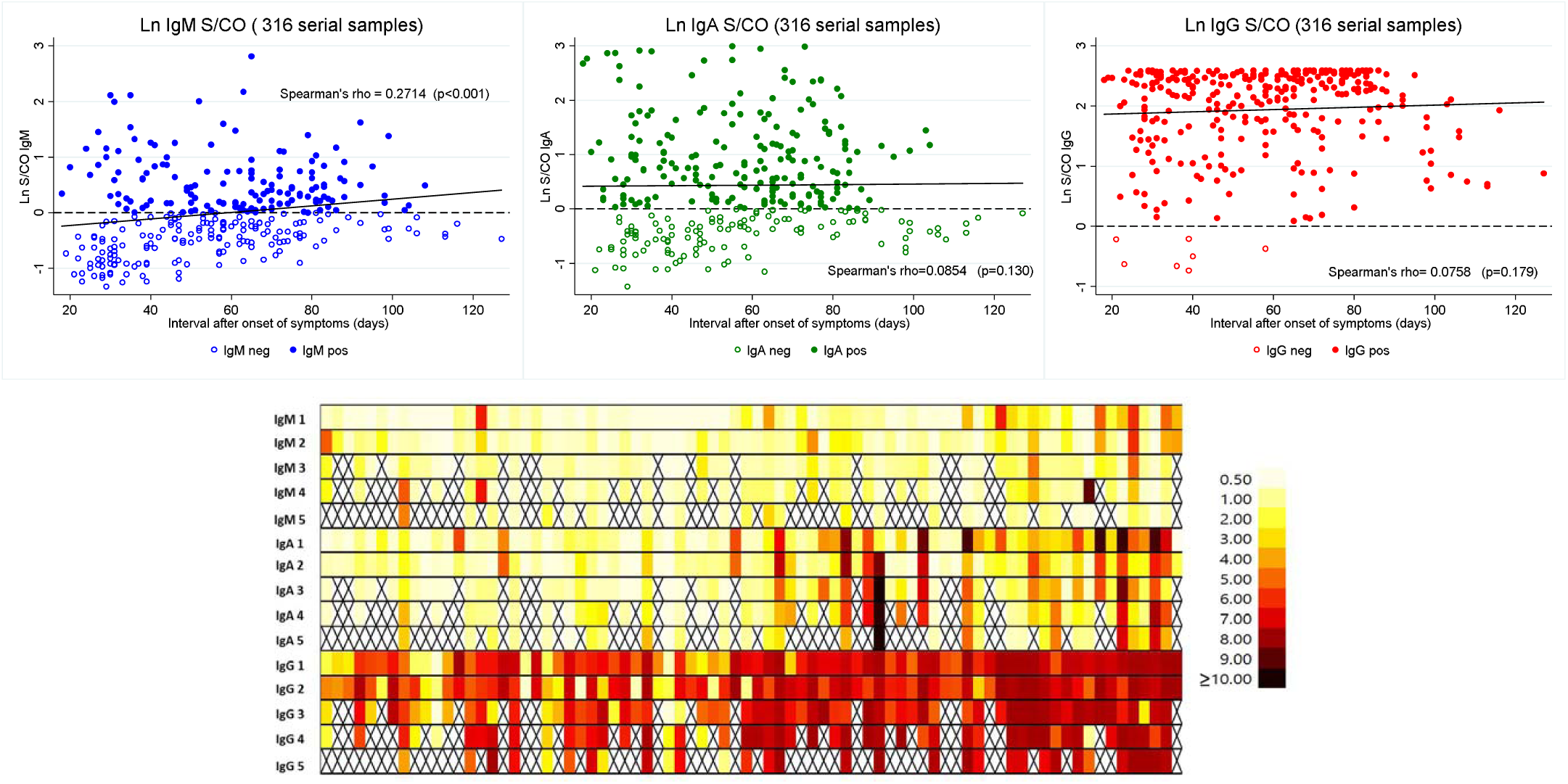
***Top:*** Anti-NP kinetics S/CO ratio of all collected samples (n=316 - transformed in natural log) for IgM (blue – left); IgA (green, middle); and IgG (red – right). The interval between collections is shown as median of days after onset of disease. Horizontal dash line represents the cut-off, measured as the natural logarithm from the S/CO ratio (cut-off= 0.2 for IgM and IgA and 0.3 for IgG). Except for IgM, there was no statistical significance for the whole group between the initial and all subsequent collections (Wilcoxon sign rank test). Hollow and solid circles represent negative and positive samples, respectively. ***Bottom***: Heat map (S/CO ratio) of all samples.

**Figure 4.**
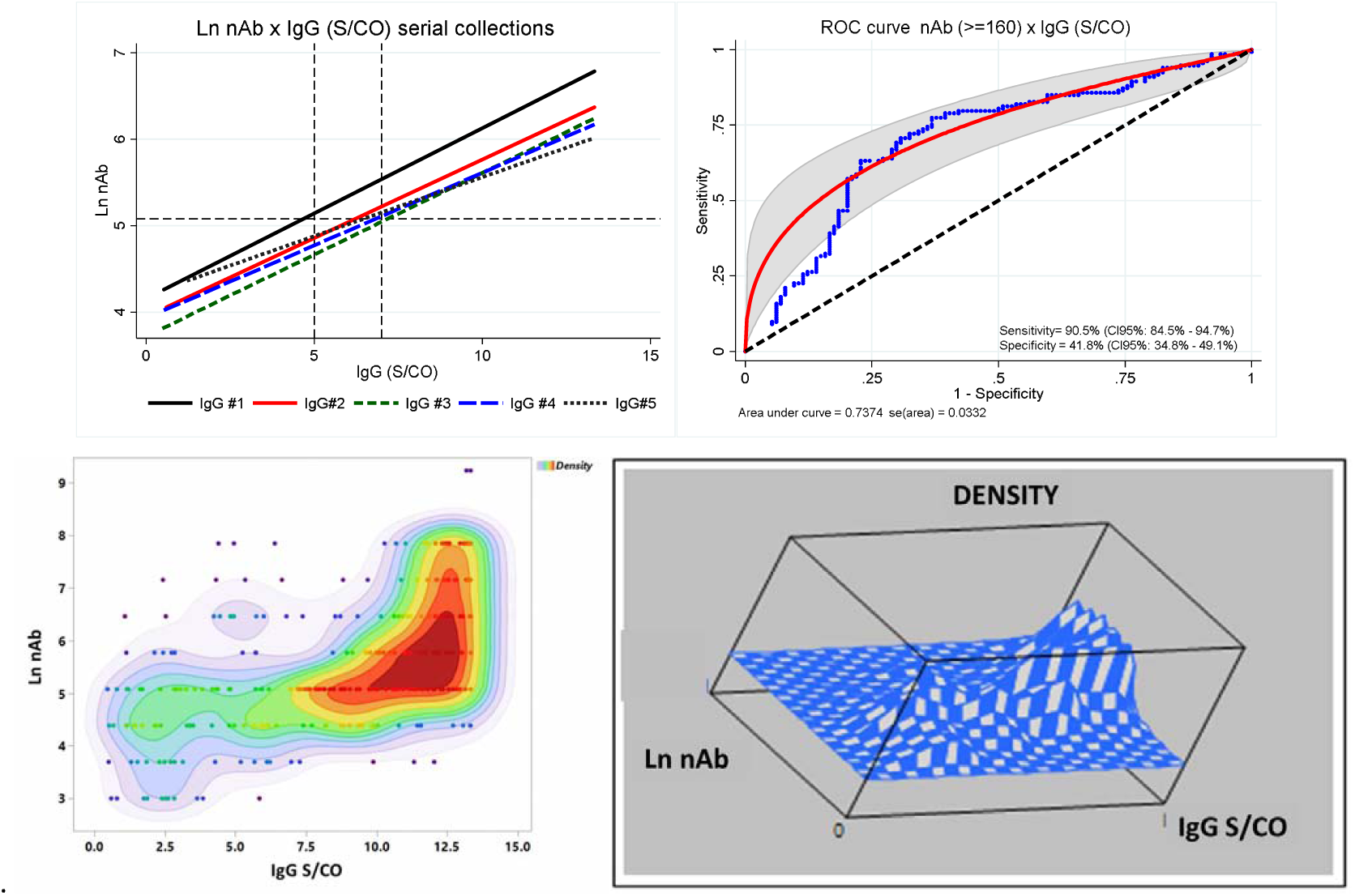
Correlation between Ln nAb and IgG (S/CO) from the cohort for five consecutive collections. ***A - Upper left***: fitted line from ordinary least square regression, showing consistent results by the corresponding adjusted R^2^ (0.3970; 0.2829; 0.3249; 0.2916 and 0.2199, respectively), with p&[lt]0.0001, except for collection #5, where p=0.001. ***B - Upper right***: ROC curve for nAb samples whose titer was ≥160. ***C - Lower left***: bivariate density contour plot showing different clusters (according to different colors). ***D - Lower right***: tridimensional mesh plot showing the corresponding peaks according to the different clusters from figure 4C.

Because a previous work has demonstrated a correlation between ABO groups and nAb titers ^21^, and due to a declining nAb titer trend, we have sought whether ABO group (particularly presence of A antigen), weight and BMI could explain this finding. The ABO antibodies titers (Anti-A, anti-B and anti-A,B, by RT/AHG) had no statistically significant difference between weight, height or BMI between cohort and control, although there was a difference for age (p=0.0212, one-sided t-test), as shown in Table 1. The cohort ABO group did not follow a random ABO type population distribution as they were not representative of the true local population (χ^2^ = 23.97, p&[lt]0.001). Despite the ABO group from controls were not balanced with the cohort’s, we thought important to have at least 25 individuals from A, B or O groups for a reasonable analysis of ABO antibodies titer distribution. Figure 5 shows no difference for ABO antibodies between cohort and controls. In addition, we stratified the ABO antibodies from the cohort based on their initial nAb titer rank (low, medium or high); except for anti-B (from A individuals), detected by the AHG test between low x medium ranks (p=0.015), we found no other statistical significance for ABO antibodies, becoming evident that ABO antibodies are not associated with maintenance of SARS-CoV-2 nAb titers.

**Figure 5.**
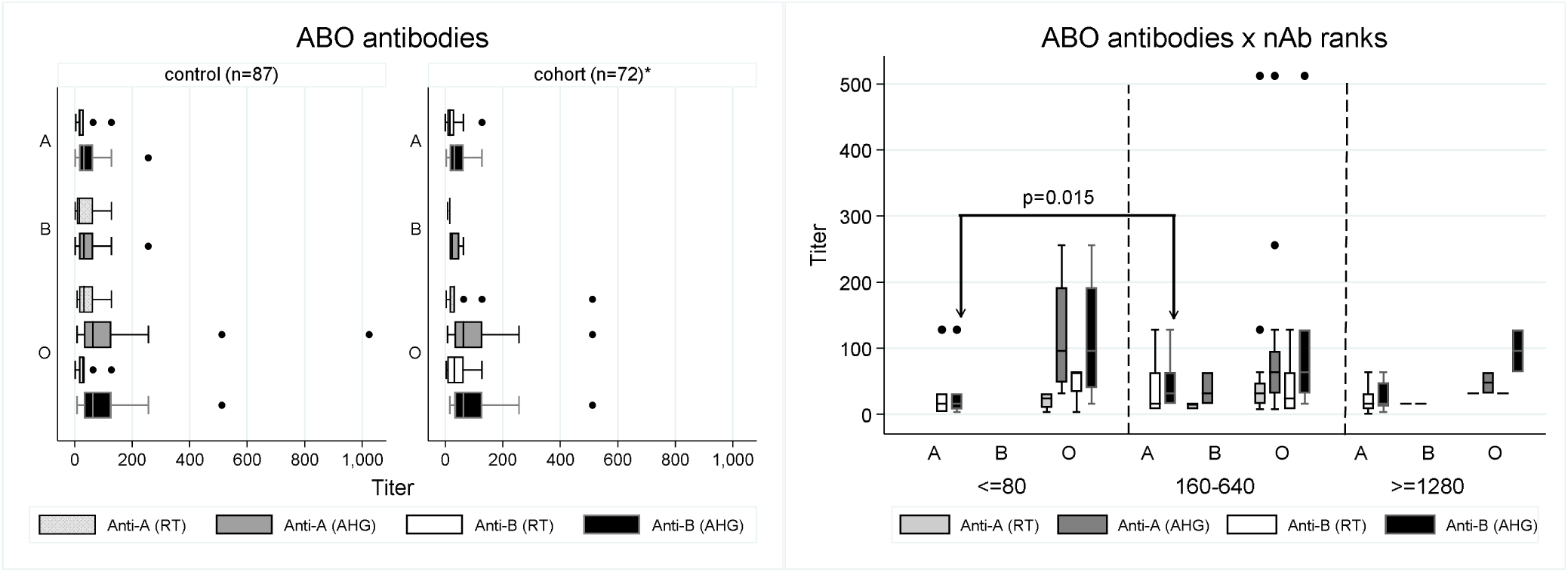
***Left***: Anti-A and Anti-B (Anti-A,B for O group) titers according to control (n=87) or cohort (n=72). * There are 6 individuals from AB group that were not included, since they do not produce anti-A or anti-B antibodies ***Right***: ABO antibody titers according to first sample nAb titer’s rank: low (nAb ≤80 - n=11); medium (640≤ nAb≥160 - n=46) and high (nAb ≥1280 - n=15) and ABO group. There was no statistical significance for ABO antibodies (anti-A, anti-B or antiA,B) either at RT or AHG between controls and cohort (Wilcoxon test); the only statistically significant titer was for anti-B (AHG) from A individuals between ranks 1 × 2 (&[lt]80 × 640≤ nAb≥160; p=0.015, Wilcoxon test).

Because of the low number of both B and AB groups, we have then merged the cases into two main blood types: those with the presence of the “A” antigen (A and AB, n= 47, 60.3%) or without it [(non-A: O and B groups; n=31 (39.7%)].We have found no association between the nAb titers (ranks), and the presence of A antigen (groups A/AB) and (p = 0.068), or absence of both A and/or B antigen (O x non-O; – 26 (33.3%) x 52 (66.7%); p = 0.087, Fisher’s exact test).

Based on BMI, individuals were classified as normal (n= 31; 39.7%), overweight (n=33; 42.3%) or obese (n=14; 18.0%); for weight, they were divided into: a) light (&[lt]75 kilos); b) medium (75-90 kilos) and c) heavy (&[gt]90 kilos). There was a good correlation between the heavy group and overweight/obese individuals (27/28 cases or 96.4%). We have previously demonstrated an important positive correlation between weight and nAb titers ^15^, also confirmed in this study. The heavier the donor, the higher nAb titer, associated with a prolonged, sustainable titer level, either during the initial sample but also for the subsequent ones, as shown in Figure 6.

**Figure 6.**
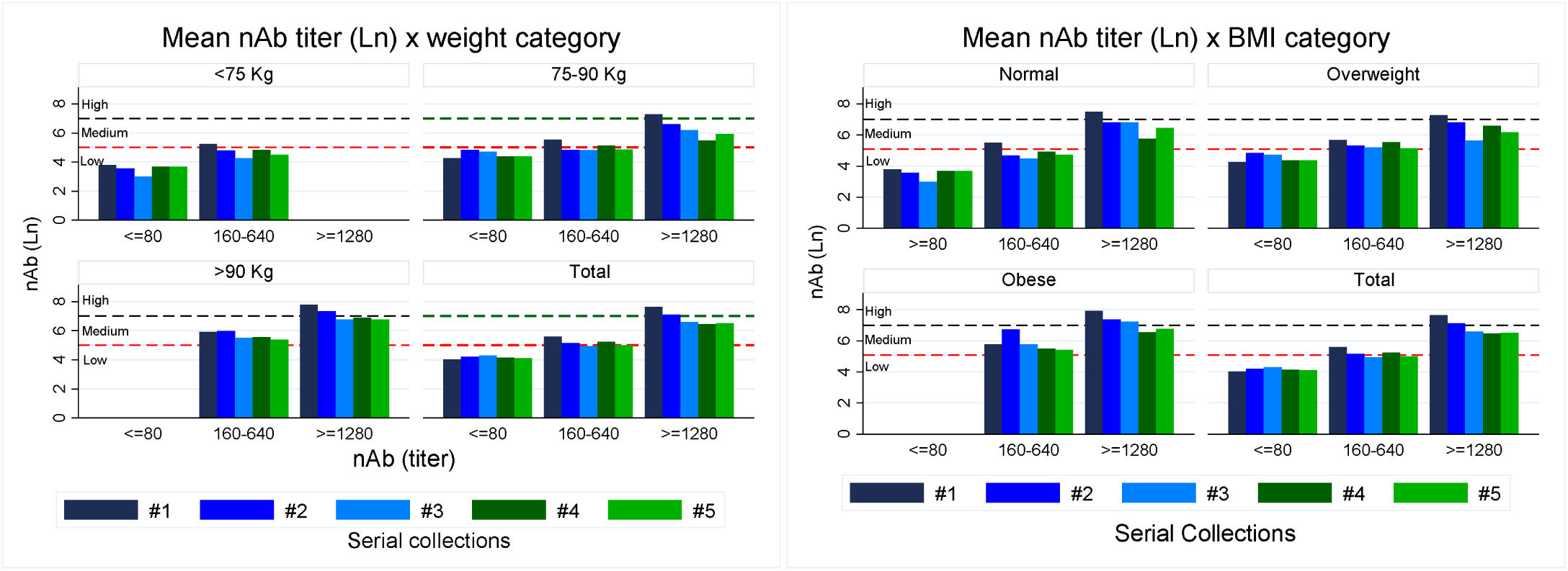
Distribution of initial neutralizing antibody (nAb) titer (Ln) based on their titer ranks (≤80: low; 160-640: medium; ≥1280: high) and cohort individual body’s weight (left): &[lt]75 (light); 75-90 (medium) and &[gt]90 kilos (heavy); or by BMI (right): normal (&[lt]25 kg/m^2^); overweight (25-29.9 kg/m^2^); obese (≥30 kg/m^2^), for the 5 consecutive collections. Horizontal dash lines represent the three limits for nAb ranks (natural logarithm). There were no cases with initial nAb titer ≥1280 in individuals weighing &[lt]75 kg; conversely, all individuals &[gt]90 kg had initial nAb titer ≥160, which persisted along the whole sample collection series. For BMI, there were some normal individuals with high nAb titers, but no obese ones had low nAb in any of the 5 collections.

Since the main objective was to investigate whether any variable could be associated with persistent high nAb titers throughout the study, we used the Kaplan Meier estimate for “non-survival”, using as final event outcome a nAb titer ≤80. The 316 collected samples spanned a total of 4820 days of study. The “survival” estimates of all samples, different ABO groups, initial nAb titer (first collection sample), and individual’s weight/BMI are shown in Figure 7. In general, nAb titers ≥160 had a median persistence of 77 days after the onset of symptoms (CI95% 62-92 days); however, only 25% remained at this level after 100 days. There was no statistical significance concerning A-type and O-type groups (log-rank χ^2^=0.65, p=0.420; HR= 1.3 - CI95%:0.7-2.5, p=0.443 and log-rank χ^2^=0.01, p=0.992; HR= 0.9 - CI95%:0.5-1.9, p=0.992, respectively). There was a major difference concerning initial nAb titers (ranks); according to crescent ranks, the median survival time varied from 29 to non-reachable (NR) data (p&[lt]0.001), the log-rank χ^2^ ranged from 7.7 to 35.1 (p&[lt]0.001) and the Hazard Ratio (HR) from 5.9 to 164.9 (p&[lt]0.001). The same was observed for weight; the median survival time ranged from 33 × 68 x 92 days after onset of symptoms (all p&[lt]0.001), the log-rank χ^2^ from 7.5 to 9.6 (p&[lt]0.01) and HR from 2.6 to 11 (p&[lt]0.01). Additional details are shown in Figure 7. It became clear that the ABO type is not important as a good predictor of high nAb titer, but weight (&[gt]90kg), BMI (overweight and obese) and the initial nAb titer (≥1280) at the first collection are the best variables for selecting CCP donors with the capacity of sustainable and adequate high nAb titers (at least ≥160) for a considerable number of days.

**Figure 7.**
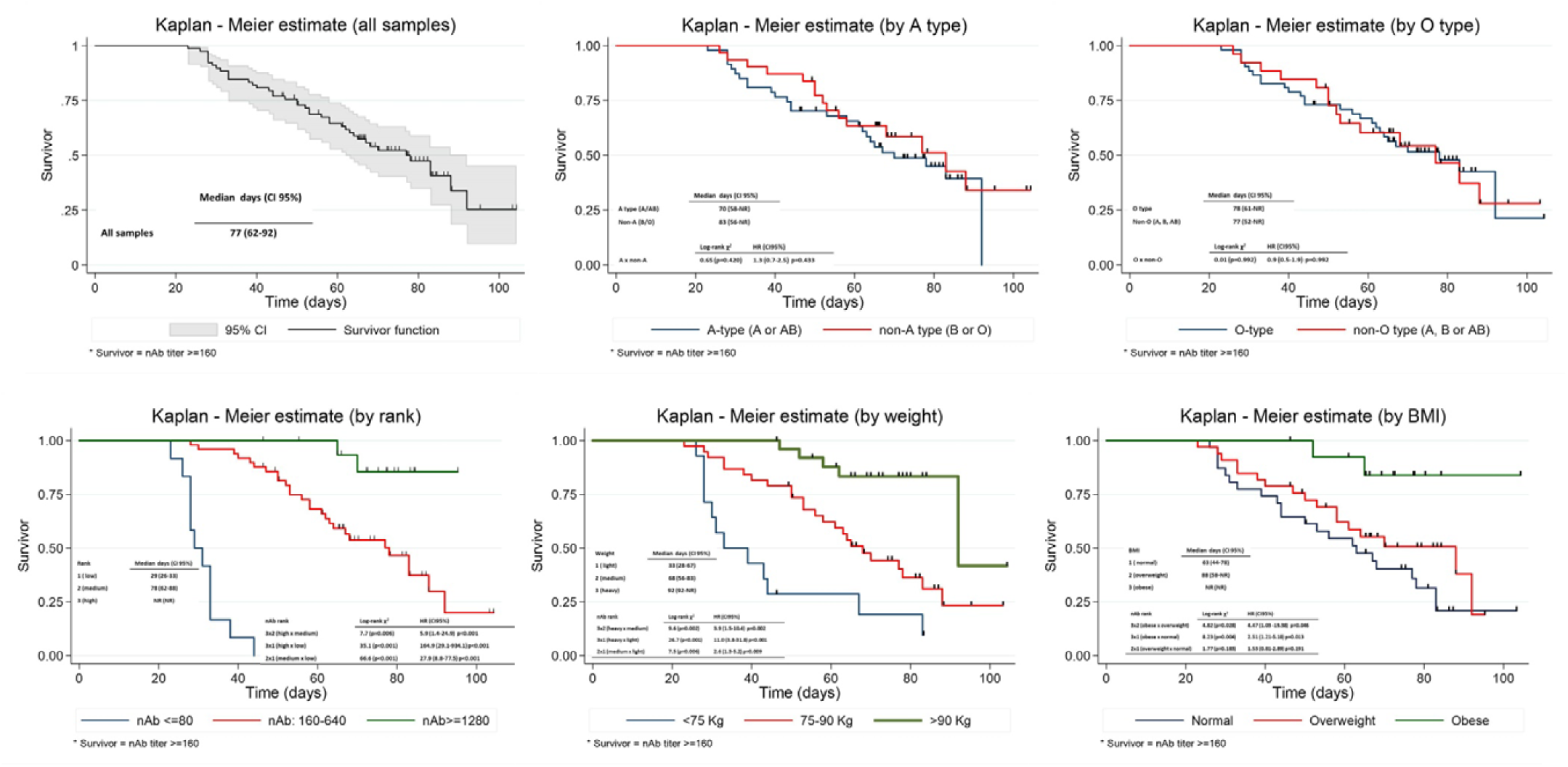
Kaplan-Meier estimate for collections that kept the nAb titers ≥160 (“survivors” – upper left) or based on: A- type (presence/absence of A antigen – A or AB/B or O; upper middle); O-type (O x non-O group; upper right); initial rank (nAb titer – lower left), weight (lower middle) or BMI (lower right). Considerable differences are observed among individuals with initial nAb titers (rank), heavy weight (kilos) or BMI (mainly obese - BMI≥30), enabling BTS to focus on special CCP donors, whose potential serial plasma collection efficiency could be more advantageous. No differences were found concerning ABO group. NR= Non reachable

## Discussion

There was no intention to evaluate the SARS-CoV-2 antibody kinetics (neutralizing or ligand antibodies) based on individual’s clinical severity, but to establish potential, simple and readily available indicators to help building a better strategy for CCP collection from a defined CCP cohort, providing CCP units with high nAb titers and maintaining them longer periods, preferably &[gt]90 days after the onset of symptoms. There are reports presenting evidences that nAbs decline to &[gt]50% of their peak level within 90 days ^15, 22^, thus, it is important to define a “golden-period” for CCP collection and extend it as longer as possible for collections. Also, there was no intention to evaluate the cohort’s immune status, neither establish a relation between the expected nAb titer decay with humoral or cellular immune impairment against COVID-19, or higher susceptibility to future re-infections.

NAbs usually start their ascending levels 10-15 days after onset of disease ^23^ with a median peak time of 33 days (IQR 24-59 days) ^24^, vanishing thereafter ^22^. So, we were assured that the initial sample collection was falling within the expected nAb peak time, as initial samples were collected after a mean period of 32.9 ± 10.3 (median = 30; range: 14-78) days.

According to Klasse ^25^, nAbs are “antibody markers of immunity against reinfection after an acute viral infection has been cleared, having the capacity to reduce viral infectivity by binding to defined viral surface particles and blocking the viral replication cycle *before* the virally encoded transcription or synthesis in the host cell”. It is yet not quite certain which is the main mechanism utilized by SARS-CoV-2 nAbs (preventing virion attachment, blocking cellular receptors, or interference with other steps necessary for cellular invasion, like endocytosis, fusion or direct penetration), nor on what moment the neutralization turns the virions infectious to non-infectious or how many molecules are necessary to achieve this effect, neither whether the existence of 2 or more different nAbs might induce an additive or synergistic effect over viral neutralization. Chen et al. ^26^ have shown that specific S1 and S2 nAbs are produced in 80.7% and 40% of cases, respectively, with 7% depending on mutual collaboration between S1 and S2-specific nAb for adequate viral neutralization. The best nAbs will be those targeted at the spike (S) protein ^27, 28^, most likely against either of subdomains on S1 (the N-terminal domain – NTD and the receptor-binding domain – RBD, responsible for viral attachment). The S2 subdomain (mediating the viral-cell membrane fusion) has not been defined at the moment as an important site targeted by nAbs. Whether neutralization escape mutants to nAbs (either naturally or passively induced) might predominate in future is unknown, although this has been recently demonstrated ^29, 30^, supporting the concept that CCP should be transfused as pooled units derived from high-tittered, potent nAb donations, lowering the probability of natural immune selection by a single antibody source (valid also for monoclonal antibodies) ^31, 32, 33^. Two groups ^14, 34^ demonstrated the isolation of potent nAb directed against the N-terminal domain (NTD), showing that both RBD and NTD might be highly immunogenic. Epitope mapping suggests that nAbs directed against the top of RBD have a stronger capacity of blocking ACE2 binding than those directed at the lateral side of RBD ^34^. It seems that nAb directed against the NTD region also plays an important role for preventing viral adhesion. It is possible that early B cell response to S protein is polyclonal and directed outside the RBD, with no nAb activity (explaining why nAbs have a peak usually at the third week of infection ^35^. A recent study has shown a good correlation between nAb titers and spike-binding antibody levels against RBD, S1 and S2 ^24^. There are compelling evidences that human CCP donations provide multiple nAb specificities, which might act on a synergistic way ^29^. However, due to the low spike amino acid sequence homology among other coronaviruses, there is a very low cross-reactivity between SARS-CoV-2 CPP against SARS-CoV-1 or MERS-CoV ^24,28,29,30,34,36,37,38^, suggesting specific viral-species inhibition. Nucleocapsid proteins (N/NP) are known to elicit antibodies earlier than S1 or S2, where most of nAbs are not targeted against these proteins. Probably anti-NP antibodies are more related to antibody-dependent cellular cytotoxicity (ADCC) viral clearance, and could be important surrogate for CCP selection of qualified donors.

There is a declining trend in nAb titers in this cohort; it seems more evident that the volunteer’s weight/BMI has a key role for maintaining higher and more sustainable titers. In addition, individuals who already presented with lower nAb titers (≤80) rarely had a rising titer up to an acceptable level in subsequent samples, and shouldn’t be considered ideal CCP donors. Some fluctuation in nAb titers was seen in the intermediate group (640≤ nAb≥160), but most of them kept levels high enough for CCP donation. On the other hand, initially high-titer individuals (≥ 1280), despite the steepest fall from the initial titer, maintained high nAb titers, placing them as the best CCP donor candidates. If this feature is combined with higher weight (&[gt]90 kilos) or overweight/obese donors, there is a great likelihood of achieving high titer donations, endured for longer periods and extending the ideal “*golden period*” CCP collections. Although this data derives from a small cohort, it provides strong evidences that selecting heavier donors increases the efficiency of CCP programs.

Our cohort comprises only from clinically moderate cases in male, young patients. This might correlate with a different nAb response observed in severe cases, recognized as having higher nAb titers and ability to inhibit *in vitro* RBD-ACE2 interaction ^26^, as far as S1 and S2-specific nAbs are concerned. Nevertheless, we have observed individuals who, despite the absence of severe symptoms have mounted a high and prolonged humoral response (persistent high nAb titers). Acute patients (especially older and male) have up to 7-8 fold higher nAb titers than asymptomatic or previously symptomatic individuals in the convalescent phase ^39,40,41,42,43,44^, maybe because of a higher viral load and subject to a longer viral exposure period. Whether higher levels equal to better protection to patients is under evaluation, since some patients who did not develop detectable nAbs were capable to recover from the infection (perhaps by other immunological pathway). One study in asymptomatic individuals ^42^ showed a lower virus-specific IgG levels than severe patients and both IgG and nAb have a higher trend to become negative at early stages (up to 40% of cases); IgG was seen in the acute phase in 81.1% and 83.8%, respectively for asymptomatic and symptomatic patients, but dropped to 60% in asymptomatic cases, while remaining positive in 87.1% of symptomatic cases. All individuals from our cohort had mild/moderate symptoms and no nAbs were seen in only 12/149 individuals (8.0%). As newer, more sensitive nAbs methods are developed (e.g, flow cytometry, sVNT) ^39, 45^, and correlated with the population seroconversion, this pre-defined minimum titer might be reviewed in the future.

The waning nAb levels observed in this study does not mean that these individuals lost their immune status or will be susceptible to re-infections in the future, as they markedly are nAb producers; only their level is below a pre-defined cut-off for donation. Even if their nAb becomes undetectable in the future, it is likely that in re-exposure, both the memory and killer T-cells will be engaged in a secondary immunologic response ^24, 46^. It is possible that CCP individuals who already presented with low nAb titers could have recovered due to additional/complementary immune mechanisms (T-cell response, cytokines, etc.) other than the classical humoral response. Recent animal models demonstrated that SARS-CoV-2 re-infection in previously immune animals was capable of induce a robust protective immune response ^30, 33, 47^, showing that immunity is maintained and is protective in subsequent viral exposures. However, human protection might last for only a period of 1-2 years ^48^.

An interesting epidemiological study from an outbreak of COVID-19 in a confined fishing vessel ^49^ demonstrated that in the only 3 people with previous nAb before the vessel’s departure, none had evidences of *bona fide* infection, whereas 103/117 individuals previously negative became infected (attack rate &[gt]85%), demonstrating the protective importance of nAbs in naïve individuals.

Since we haven’t studied female volunteers, we cannot confirm whether there is a gender difference in nAb titers, as described by others ^24^, although gender difference should be adjusted for weight or BMI. Age is also an important factor, but given that our cohort comprises mainly younger individuals (36.7 ± 9.1, median =36.5 years), we couldn’t find any age trend.

We didn’t find any significant change on IgG and IgA levels during the whole study period (Figures 3), except for a minor rise on IgM levels, in contrast with previous data ^15,42,50^, where there was a drop in IgG and IgM levels after the early convalescent period or the time between onset of symptoms and first sample collection. Both IgG1 and IgG3 directed against S1, NP, RBD and ECD (ectodomain) are detected as early as 4-7 days after infection; it seems that IgG1 is the predominant nAb class ^34^ and remains stable for at least 90 days ^39^, whereas IgA against NP and S1 is hardly detectable after this period ^51^ .The persistent correlation between IgG anti-NP S/CO levels and nAb titer confirms our initial findings ^15^ and provides additional evidences that this test is a good surrogate screening for high-tittered CCP, with well-defined clusters and good sensitivity, evidenced by the ROC analysis (Figure 4).

The association between ABO group and viral infections (HIV, measles, hepatitis B and SARS) has already been described ^52, 53, 54, 55, 56^. Zhao *et al*. ^57^ showed higher odds for A blood group COVID-19 cases, also observed for SARS ^58^. In New York, a preliminary study showed higher odds in group A and B versus group O, not explained by associated risks, but with no association with intubation or death rate. The genomewide association study (GWAS) ^16^ evaluated more than 8,5 million single-nucleotide polymorphisms and identified the rs657152 polymorphism (locus 9q34.2) related with *ABO* blood group locus, which showed a higher risk in blood group A than other blood groups (OR= 1.45; P= 0. 0001) and a protective effect in blood group O (OR= 0.65; P= 0.000001). A French study ^59^ demonstrated that seropositivity among blood donors is higher in group A than O donors (3.86% x 1.32%, p=0.014). It is possible that ABO group is not related to a predisposing risk of being infected by SARS-CoV-2, but to a different predisposition of COVID-19 severity ^60^. A meta-analysis evaluating seven studies with 13 subgroups, with &[gt]7500 cases and &[gt]2.9 million controls ^61^ indicates that SARS-CoV-2 positive individuals are more likely to be group A (OR=1.23; 95%CI: 1.09-1.40), whereas group O individuals are more protected (OR=0.77; 95%CI: 0.67-0.88), with no evidences for groups B or AB.

Whether the risk is associated with the presence/absence of A antigen, or the corresponding antibody, i.e, the presence/absence of anti-A from O/B or A/AB groups, respectively due to inhibition of SARS-CoV-1 to adhesion of spike protein to ACE2 receptor in the early stages of infection is debatable; ^62, 63, 64^. The spike protein possesses several N-linked glycosylation sites, decorated by A or B glycans in A, B or AB individuals; these carbohydrate epitopes act like a shield surrounding the RBD^Error! Reference source not found., 66, 67^, whose cellular adhesion could be hampered by the presence of A or B antibodies, blocking the interaction between SARS-CoV-1 S-protein and the host ACE2.

Arend ^68^ proposed a different model for SARS-CoV-2 invasion, initially independently from ABO group, via the formation of a hybrid A-like/Tn (T-nouvelle) structure (different from the specific blood group A epitope), acting as a functional molecular bridge between the spike protein and host mobilization (via TMPRSS2), allowing the final viral invasion into the host’s cell (initial invasion could be present both in group O and non-O individuals, via the host’s GalNac metabolism pathway). After invasion, SARS-CoV-2 newly formed virions from group O individuals replace this initial metabolic pathway by the mucin-type fucosylation and consequent synthesis of hybrid H-type antigen, being unaffected by the innate anti-A/Tn or anti-B/Tn isoagglutinins, leading to a secondary IgG response; also, viral binding is reduced, because it happens only via the hybrid-H type antigen, since the initial anti-A or -B/Tn isoagglutinins are preserved, possibly explaining the protector effect on group O individuals. On the other hand, group A, B and AB hosts replace the intermediate A-like/Tn binding site by hybrid A or B allelic mucin types, with additional “physiological downregulation” of anti-A or –B/Tn activities, with consequent decrease of anti-A or anti-B isoagglutinin levels and/or clonal selection, resulting in higher contact between the pathogen and host, leading to a lower protective status in A, B or AB hosts. This newly proposed mechanism must be confirmed by additional studies but characterizes an intriguing mechanism of viral invasion and innate natural protection. Anyway, this explanation seems plausible for the protector effect observed on O group individuals ^62^, and a schematic model of viral neutralization capacity based on ABO groups from potential naïve individuals is shown in Table 3. Either by hybridization of ABO group molecules or by mimicking the related metabolic pathways, SARS-CoV-2 might, hypothetically evade human immunity until other stronger immune mechanisms (humoral, cellular) are present. Whether anti-A,B from O individuals (with predominance of IgG class) are more protective than anti-A from B (mainly IgM class) is still debatable ^62^.

**Table 3.** Theoretical viral neutralization capacity based on ABO groups from potential naïve individuals, according to the viral ABO phenotype from the infected spreader. * Only Bombay individuals could, in theory be more resistant to SARS-CoV-2 from a group O spread source (natural presence of anti-H). Due to its extremely rare condition, it will be unlikely to have enough cases to confer a statistically valid conclusion in this group. Light grey cells denote the same isotype virions between spreader and newly infected individual (adapted from Breiman) ^Error! Reference source not found^.

| Spreader |  | Potential viral neutralization capacity from naïve infected individuals, based on their ABO phenotype |  |  |  |
| --- | --- | --- | --- | --- | --- |
| Virus phenotype | Viral spike glycans | O * | B | A | AB |
| O | H | ↑↑↑ | ↑↑ | ↑↑ | ↑↑ |
| B | H, B | ↓↓ | ↑↑↑ | ↓ | ↑↑ |
| A | H, A | ↓↓ | ↓ | ↑↑↑ | ↑↑ |
| AB | H, A, B | ↓↓ | ↓ | ↓ | ↑↑↑ |

Although ABO antibodies could play a protective role for SARS CoV-2 infection, one needs to explain why A groups have a trend to produce higher nAb levels, as opposed to O individuals, at least in early stages ^21^. It is possible that O individuals, who naturally have anti-A antibodies and might have a lower viral adhesion, could reduce their antigenic immune stimulus, leading to a lower nAb production, whereas the proposed higher viral adhesion in A, B and AB infected individuals might explain their higher nAb titers.

A natural inhibition by anti-A, B or A,B antibodies with consequent reduced cell invasion might prevent opsonization into host cells and subsequent neutralization through the complement system, potentially leading to a lower cellular immune response, with decreased cytotoxic T-cell production, explaining the lower risk of severe clinical effects in O infected patients. It is also likely that either the anti-A, -B or -A,B isoagglutinin class and their corresponding titers might also exert different initial neutralization role; Focosi ^69^ suggests that only anti-A IgG, with titers &[gt]1:16 (more common in group O individuals) might benefit from this mechanism. However, once SARS-CoV-2 overcomes the initial natural neutralization barrier and infects recipient’s cells, then the newly formed viruses will bear the corresponding recipient’s spike glycans and the next viral generation will not be subject to the same initial ABO neutralization for attachment and entry, due to lack of ABO viral glycan’s incompatibility. After this stage, other immune pathways will take the lead in controlling viral infection. This might explain why, once the full viral infection cycle has been established, there is not a striking difference on clinical symptoms on patients according to their ABO phenotypes ^57,64,70^. Other cofounders related to ABO polymorphism have also been detected (C3 and ACE1), possibly exerting additional activities ^71^.

Despite we have found initial association between A-type donors and higher nAb levels, unfortunately this variable hasn’t sustained significant correlation after few weeks of study.

Obesity is defined by WHO when BMI is ≥ 30kg/m^2^ ^72^, associated with visceral adipose tissue expansion, inflammation by secretion of pro-inflammatory cytokines (TNF-α, IL-6), adipokines (leptin) and reduction of the anti-inflammatory adiponectin ^73, 74^, leading to a pro-inflammatory status, major oxidative stress and impaired immune response. Other co-morbidities are associated, some also with COVID-19 severity (diabetes and hypertension)^75,76^. The renin-angiotensin-aldosterone system (RAAS) is recognized as having two opposite counter-regulatory arms and several molecules, whose function is to regulate the fluid balance homeostasis, blood pressure and cardiorenal function. A deeper explanation of RAAS in normal or pathologic status can be found elsewhere ^77,78,79,80^. The most important factor between COVID-19 and RAAS is the angiotensin-converting-enzyme 2 (ACE2), which converts AngI to Ang (1-9) and AngII to Ang(1-7), acting as the cellular receptor for SARS-CoV-2 (also SARS-CoV and NL63), via the RBD, whose affinity is 10-20-fold of SARS-CoV; ACE2 is mostly bound on cellular membranes (soluble form in minute amounts in the bloodstream). The cellular Ang (1–7) receptor Mas is able to reduce the pro-inflammatory cytokine release. Since obesity is associated with a reduced expression of the ACE2/Mas axis and overactivation of RAAS (also related to more severe SARS-CoV-2 cellular infection) and prolonged viral shedding at the adipose tissue, with excessive chronic cytokine release ^81^, it is plausible to consider that this mechanism might explain the higher and prolonged production of nAbs, via a downregulation of ACE2/Mas function ^82,83^ and higher inflammatory activity. It is possible that other genetically-induced mechanisms (e.g. HLA, reduction of cytotoxic activity of NK cells, reduced ADCC, imbalance between the pro-inflammatory leptin and the anti-inflammatory adiponectin) might play additional role in this effect.

We recognize as a weakness that this analysis did not include women, thus we cannot assure the same nAb behavior. However, the normal lower weight and adiposity, combined with a lower adipocyte ACE2 response in regards to the RAS, also controlled by estrogens in females ^81^, might produce a different response, necessary to be confirmed/refuted by studies with female CCP donors. In addition, this population is subject to a selection bias; the cohort obese male individuals are healthy, not representing the general obese population.

In conclusion, despite the real therapeutic value of CCP for COVID-19 remains uncertain ^3^, we have found that both the initial high nAb titer, particularly if ≥ 1280, heavy donors (&[gt]90 kilos), overweight or obese individuals (BMI ≥25) could bear high nAb titers for a prolonged period of time (&[gt]100 days), enabling BTS to optimize their CCP recruitment scheme and reducing the number of low titer CCP units. Naturally, we cannot define what would be the nAb behavior after 100 days, demanding additional studies; however, we consider that defining strategies to target at special donors, able to donate for at least 100 days is a reasonable contribution to improve the efficiency of CCP programs.

## Data Availability

I hereby state that all data will be availabe under special request

## Acknowledgements

Authors would like to thank Márcia Milena Pivato Serra, PhD, Assistant Professor, Statistics Department, University of Campinas (UNICAMP) for the statistical suggestions and analysis, and JoaoRoberto de Sá, MD, PhD, Associated Professor – Discipline of Endocrinology and Metabology, Federal University of SaoPaulo (EPM/UNIFESP), for comments concerning obesity and COVID-19. We would also like to thank all participants in this study, who agreed on providing serial samples, even after recovering from this newly pandemic viral disease.

## Author contribution

Conceptualization: SW, RFW, RMF; investigation: SW, RFW, RMF, GC, PS, RA, MAB, RRGM, DA, CPS, ED.; formal analysis: SW; resources: LFLR, AAC, MA.; writing: SW. This project was partially supported by the initiative “Todos Pela Saude”.

