## Supplement Tables and Figures for "Correlation of Body Mass Index (BMI), initial neutralizing antibodies (nAb), ABO group and kinetics of nAb and anti-nucleocapsid (NP) SARS-CoV-2 antibodies in convalescent plasma (CCP) donors – A longitudinal study with proposals for better quality of CCP collections"

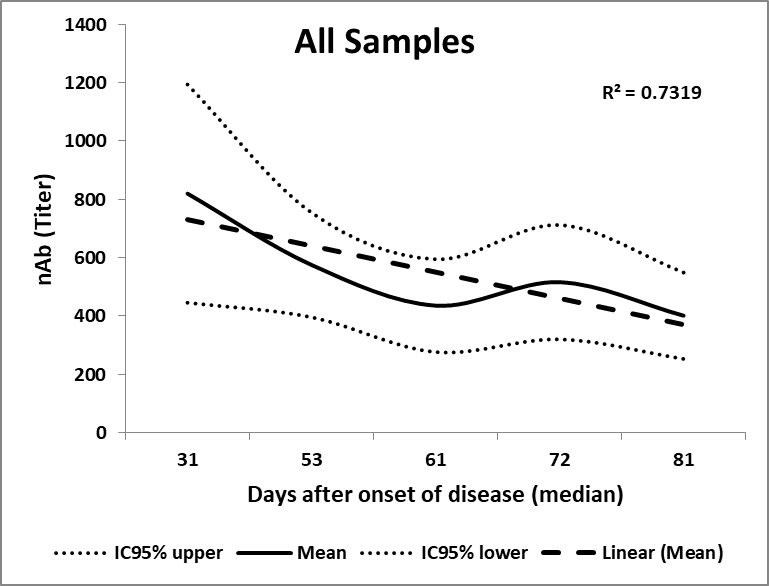

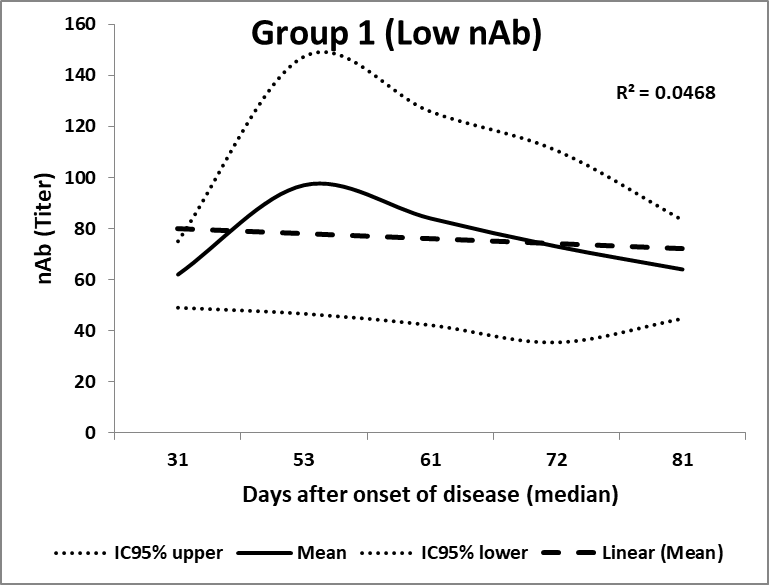

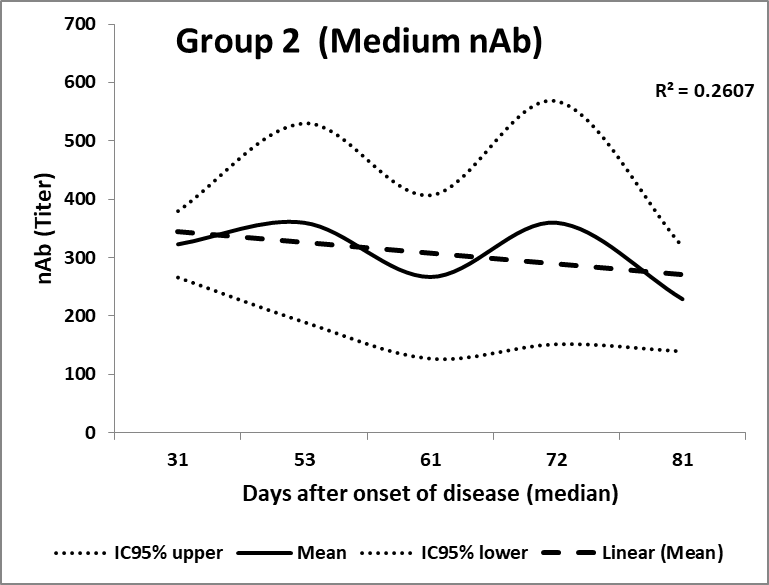

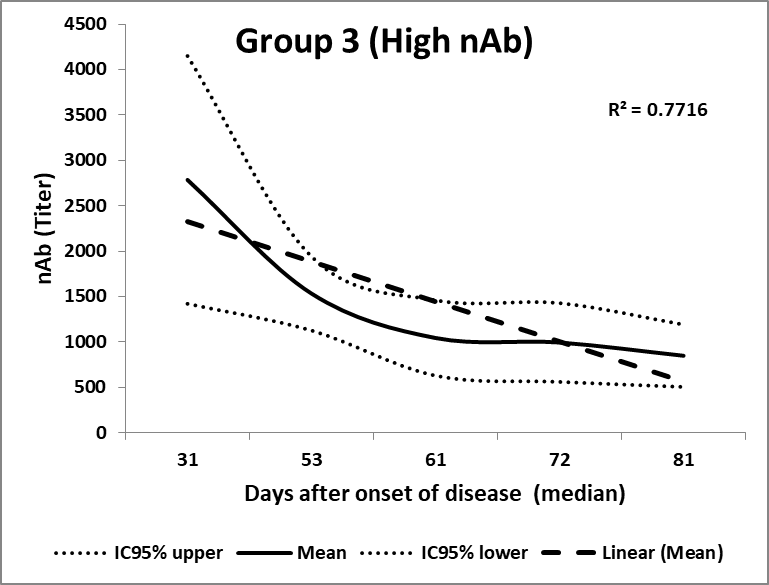

Figure S1 – Neutralizing antibodies (nAb) of cohort (n=78 individuals; 316 collections), showing a progressive decline in nAb titers (all cohort – upper left quadrant), but with different patterns based on their initial nAb titer (ranks). Group 1 (low; n=12) kept a sustained low nAb titer, which prevented the participants from future plasma donation. Group 2 (medium; n=49) had a modest decline in nAb titers, though most of participants sustained levels high enough for donation. Group 3 (high, n=17), despite showing the most persistent nAb titer decline, always kept their levels above the minimum necessary for plasma donation (&[ge]160), being the best candidates for a CCP program. The interval between collections is shown as median of days after onset of disease.

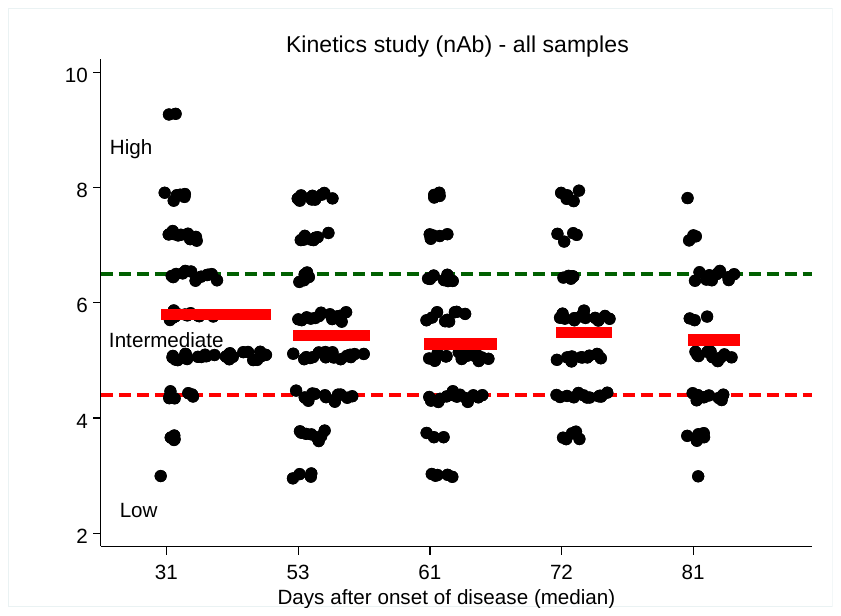

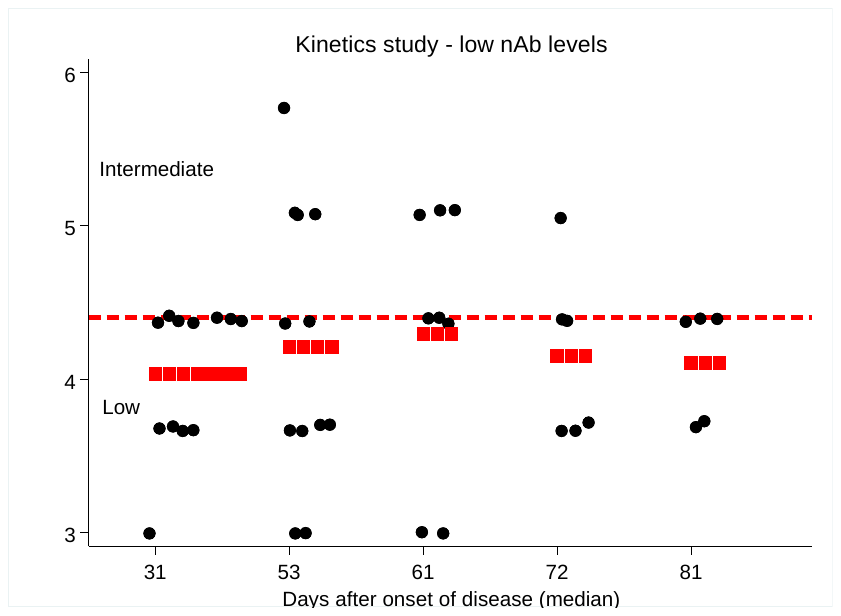

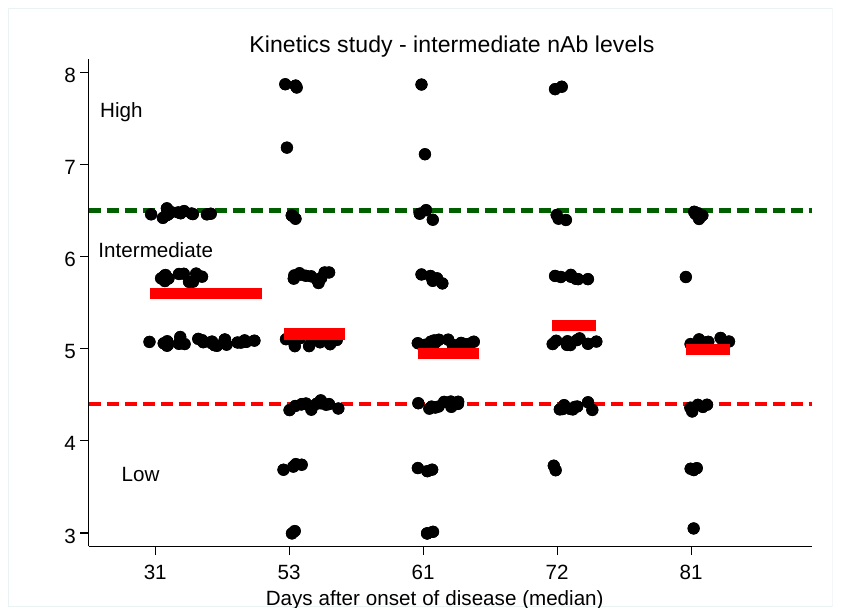

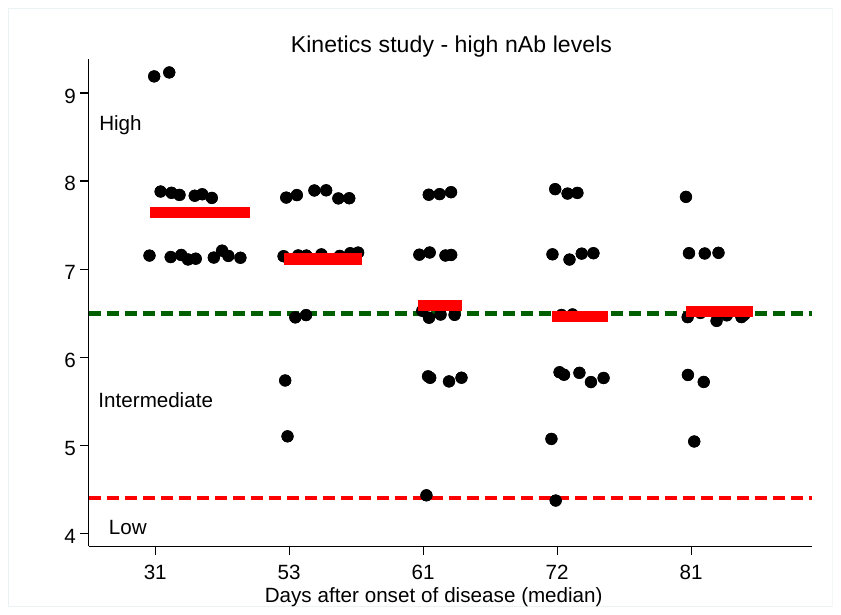

Figure S2 – Neutralizing antibodies (nAb) titers of samples based on their initial rank classification: all samples (n=78, upper left); group 1 (low, nAb &[le]80 - upper right; n=12); group 2 (intermediate, 640&[le] nAb&[ge]160 - lower left; n=49); group 3 (high, nAb &[ge]1280, lower right; n=17). The interval between collections is shown as median of days after onset of disease. Horizontal dash lines represent the three limits for nAb ranks (transformed into natural logarithm). Statistical significance for the whole group was observed for both the initial nAb (titer) and all subsequent collections, and also between nAb collection #2 x collections #3 and #5among serial samples (all p&[lt]0.02; Wilcoxon sign rank test).

|  | Control (n=87) | | | Cohort (n=78) | |
| --- | --- | --- | --- | --- | --- |
| Blood Group | N | % |  | N | % |
| A | 30 | 34.5% |  | 41 | 52.6% |
| AB | NA |  |  | 6 | 7.7% |
| B | 27 | 31.0% |  | 5 | 6.4% |
| O | 30 | 34.5% |  | 26 | 33.3% |

Table S1 – ABO groups from controls and cohort. No AB group was included, as they normally do not produce ABO antibodies.
